# Acetyl-L-leucine for Niemann-Pick type C – a multi-national, rater-blinded phase II trial

**DOI:** 10.1101/2020.12.22.20248704

**Authors:** Tatiana Bremova-Ertl, Jen Claassen, Tomas Foltan, Jordi Gascón Bayarri, Paul Gissen, Andreas Hahn, Anhar Hassan, Anita Hennig, Simon Jones, Miriam Kolniková, Kyriakos Martakis, Uma Ramaswami, Reena Sharma, Susanne Schneider

## Abstract

**Background:** Niemann-Pick disease type C (NPC) is a rare autosomal recessive neurodegenerative disorder characterized by symptoms such as progressive cerebellar ataxia and cognitive decline. The modified amino acid N-acetyl-leucine has been associated with positive symptomatic and neuroprotective, disease-modifying effects in observational case studies and animal and cellular models of NPC. Therefore, the effects of the active L-enantiomer, N-acetyl-L-leucine (NALL, Sponsor Code IB1001) were evaluated in paediatric and adult patients with NPC.

**Methods:** We conducted a 9-center, multinational, open-label, rater-blinded Phase 2 study to assess the safety and efficacy of NALL for the treatment of pediatric (≥ 6 years) and adult patients with NPC (IB1001-201 clinical trial). Eligible patients were assessed during three study phases: a baseline period (with or without a study run-in), a 6-week treatment period (dosage of NALL 4 g/d in patients aged ≥13 years; weight tiered doses for patients aged 6-12 years based on approximately 0.1 g/kg/day), and a 6-week post-treatment washout period. The primary outcome was based on the Clinical Impression of Change in Severity (CI-CS) assessment (assessed on a 7-point Likert scale) performed by blinded, centralized raters who compared videos of patients performing a pre-defined primary anchor test (either the 8-Meter Walk Test (8MWT) or 9-Hole Peg Test of the Dominant Hand (9HPT-D)) at different study periods. Secondary outcomes included the cerebellar function evaluations, namely the Scale for Assessment and Rating of Ataxia (SARA), the Spinocerebellar Ataxia Functional Index (SCAFI), and the Clinical Global Impression Scales (CGI) as well as the EuroQol-5D/VAS.

**Results:** Thirty-three subjects aged 7 to 64 years with a confirmed diagnosis of NPC were enrolled according to the trial protocol between 04 September 2019 and 30 January 2020. Thirty-two patients were included in the modified intention-to-treat analysis. IB1001 met its CI-CS primary endpoint (mean difference 0.86 ((90% CI 0.25,1.75, *p*=0.029). IB1001 also met secondary endpoints, including improvement during treatment on the SARA scale (mean difference −1.19 (90% CI −1.8, −0.5, *p*=0.001)), investigator’s CGI-C assessment (mean difference from baseline to the end of treatment 0.6 (90% CI 0.5, 1.0, *p*<0.001)), clinically worsening over the washout period on the SARA (mean difference 1.45 (90% CI 0.5, 2.0, *p*=0.002)) and investigator’s CGI-C assessment (washout mean difference −0.5 (90% CI −1.0, 0.0, *p*=0.006). IB1001 was well-tolerated with no treatment related serious adverse reactions occurring.

**Conclusions:** Consistent with its pharmacological action, IB1001 rapidly improved symptoms, functioning, and quality of life in 6-weeks, the clinical effect of which was lost after the 6-week, post-treatment washout period. High consistency and statistical significance between the primary and secondary endpoints demonstrate a clear, clinically meaningful improvement with IB1001. IB1001 was well-tolerated and no drug-related serious adverse events were reported, demonstrating a favorable risk/benefit profile for the treatment of NPC (Funded by IntraBio; ClinicalTrials.gov number, NCT03759639; EudraCT number 2018-004331-71).

## BACKGROUND

Niemann-Pick disease type C (NPC) is an rare (incidence 1:100,000), prematurely fatal, autosomal recessive, neurovisceral lysosomal storage disease that predominantly affects children. Adolescent and adult onset cases are being increasingly recognized (Geberhiwot et al., 2018). The disease typically begins in early childhood, is chronic, progressive, and severely reduces quality of life. The presentation of NPC is characterized by broad heterogeneity in systemic, psychiatric, and neurological symptoms, which depends on age of onset of neurological symptoms (Vanier and Millat, 2003). There is broad inter-individual phenotypic variability, including of age at onset and rate of progression. This renders an assembly of well-matched cohorts of NPC patients for controlled trials difficult to achieve.

Treatment of NPC is currently limited to reducing the rate of disease progression with the substrate reduction therapy drug miglustat (Zavesca™), which is approved in the European Union and several other countries, but not in the United States.

N-acetyl-L-leucine (NALL) is the L-enantiomer of N-acetyl-DL-leucine, a modified, acetylated derivative of a natural essential amino acid (Leucine) that has been available in France since 1957 as a racemate (equal amounts of both D- and L-enantiomers) under the trade name Tanganil™ (Pierre Fabre Laboratories, France) as a treatment for acute vertigo. Prior observational studies assessing the effect of N-acetyl-DL-leucine in patients with NPC suggest a beneficial symptomatic, and neuroprotective, disease modifying effect of this agent. In a case series, short-term treatment with N-acetyl-DL-leucine was found to improve ataxia, cognition, and quality of life in 12 patients with NPC (Bremova et al., 2015). Subsequent long-term case series and pre-clinical studies demonstrated the neuroprotective, disease modifying effect of treatment in NPC (Cortina-Borja et al., 2018; Kaya et al., 2020a). In all, the compound was well tolerated with no serious side effects.

Animal studies in the NPC mouse model have shown that the L-enantiomer, i.e. NALL, has potential clinical benefits compared to the racemic mixture. One mechanism of action of NALL is the activation of cerebral glucose metabolism in the cerebellum, correlated with enhanced cerebellar activity (Günther et al., 2015; Tighilet et al., 2015). In an animal model of NPC, N-acetyl-DL-leucine and its enantiomers significantly reduced ataxia in *Npc1*^*-/-*^ mice, when commenced symptomatically (from 8-9-weeks of age) and pre-symptomatically (from 3-weeks of age) (Kaya et al., 2020a). These studies specifically identify the L-enantiomer as the neuroprotective isomer, observed to significantly delay the onset of functional decline (gait abnormalities, motor dysfunction), the decline in general health and condition, as well as to slow disease progression and prolong survival (whereas the D-enantiomer did not). Similar effects of NALL were found in an analogous animal model of another lysosomal storage disease, the Sandhoff mouse (Kaya et al., 2020b). It is important to note that the dosage used in these *in vivo* studies (0.1 g per kg per day) is roughly equivalent to the dose used in previous observational clinical studies with the racemate and the IB1001-201 clinical trial.

Finally, pharmacokinetic studies suggest that the D-enantiomer could accumulate relative to the L-enantiomer during chronic administration of the racemate, which has the potential for long-term negative effects (Churchill et al., 2020). Therefore, in this clinical trial, the effects of NALL were evaluated.

## METHODS

### Study Design

The IB1001-201 clinical trial was one of three multinational, open-label, rater-blinded trials which utilize a single master protocol to investigate NALL (IB1001) for the treatment of three rare, neurodegenerative diseases (in addition to NPC, GM2 Gangliosidosis [NCT03759665] and Ataxia-Telangiectasia [NCT03759678]). This IB1001 master protocol was designed through a collaboration between National Regulatory Agencies, leading clinical experts, patient organizations, and the industry sponsor to address of the unique ethical and practical challenges to conducting clinical trials for these orphan, heterogenous patient populations. Details of the trial methods, rational, design, and oversight have been previously published (Fields et al., 2020, preprint).

The IB1001-201 clinical trial was separated into two study phases in order to enable the investigation of both the symptomatic (“Parent Study”), and long-term (“Extension Phase”) safety and efficacy of NALL. The results of the Parent Study are reported below.

### Study Procedures and Treatment

Adult and pediatric subjects with a confirmed diagnosis per the current recommendations for the detection and diagnosis of NPC were (Patterson et al., 2017) recruited at 9 clinical trial sites in five countries (Germany, Slovakia, Spain, the United States and United Kingdom). The Parent Study consisted of three study periods: a baseline period (with or without a study run-in washout from prohibited medications), a treatment period and a post-treatment washout period, with two patient visits per period (**Figure 1**). Duration of the treatment phase and washout phase were 42 days (+7 days) each, per protocol. This study was ongoing during the Coronavirus Pandemic (COVID-19), which significantly impacted the schedule of events.

**Figure 1.**
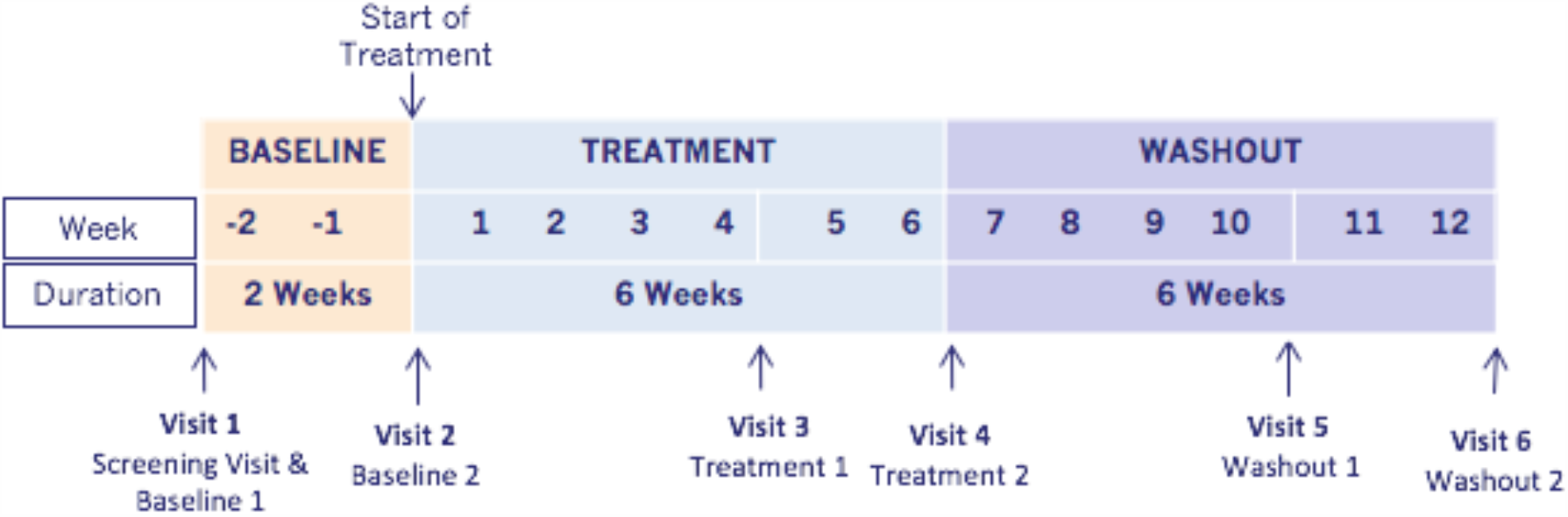
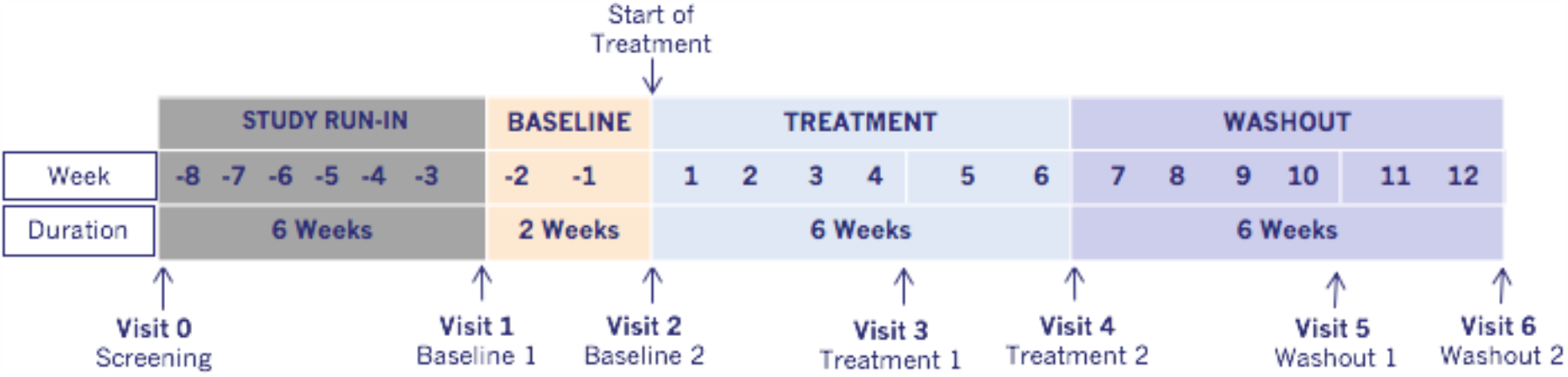
Study Schemas for the IB1001-201 clinical trial. **A)**. Schema for naïve patients. **B)**. Schema for non-naïve patients.

At the initial screening visit, patients were classified as either “naïve” or “non-naïve” depending on their use of prohibited medications (see below) within the past 6 weeks. “Non-naïve” patients were given the one-time opportunity to undergo a minimum of 42 days washout before returning for the baseline 1 visit.

During the treatment period, patients aged ≥13 years or aged 6-12 years weighing ≥35 kg received 4 g/day of orally administered IB1001 (2 g in the morning, 1 g in the afternoon, and 1 g in the evening). Patients aged 6-12 years weighing <35 kg received weight tiered doses based on approximately 0.1 g/kg/day.

After their final visit of the Parent Study (Visit 6), patients were given the opportunity to enter an extension, open-label study lasting 12 months which is still ongoing.

### Study Population

The inclusion criteria are given in Table 1.

**Table 1.** Inclusion Exclusion Criteria

| Inclusion Criteria | Exclusion Criteria |
| --- | --- |
| Individuals who meet all of the following criteria are eligible to participate in the study:<br>1. Written informed consent signed by the patient and/or their legal representative/ parent/ impartial witness<br>2. **IB1001-201 (NPC) EU:** Male or female aged $\geq 6$ years in Europe OR $\geq 18$ years in the United States with a confirmed diagnosis of NPC at the time of signing informed consent. Confirmed diagnosis includes<br>a) Clinical features and positive biomarker screen and/or filipin test without genetic tests results (has not been performed)<br>b) Clinical features and positive genetic test<br>c) Clinical features and positive biomarker screen and/or filipin test but only one NPC mutation identified on genetic test<br>d) Clinical features with positive biomarker screen and/or filipin test and positive genetic test<br>**IB1001-201 (NPC) US:** Male or female aged $\geq 6$ years in Europe OR $\geq 18$ years in the United States with a confirmed diagnosis of NPC at the time of signing informed consent. Patients must have clinical features of NPC and a positive genetic test for mutations in both copies of NPC1 or in both copies of NPC2. | Individuals who meet any of the following criteria are not eligible to participate in the study:<br>1. Asymptomatic patients<br>2. Patient has clinical features of NPC and a positive biomarker screen and/or filipin test, but a completely negative result on a previous genetic test for NPC<br>3. Patients who have any of the following:<br>a) Chronic diarrhea;<br>b) Unexplained visual loss;<br>c) Malignancies;<br>d) Insulin-dependent diabetes mellitus.<br>e) Known history of hypersensitivity to the Acetyl-Leucine (DL-, L-, D-) or derivatives.<br>f) History of known hypersensitivity to excipients of Ora-Blend® (namely sucrose, sorbitol, cellulose, carboxymethylcellulose, xanthan gum, carrageenan, dimethicone, methylparaben, and potassium sorbate).<br>4. Simultaneous participation in another clinical study or participation in any clinical study involving administration of an investigational medicinal product (IMP; ‘study drug’) within 6 weeks prior to **Visit 1**.<br>5. Patients with a physical or psychiatric condition which, at the investigator’s discretion, may put the patient at risk, may confound the study results, or may interfere with the patient’s participation in the clinical study.<br>6. Known clinically significant (at the discretion of the investigator) laboratories in hematology, coagulation, clinical chemistry, or urinalysis, including, but not limited to:<br>a. Aspartate aminotransferase (AST) or alanine aminotransferase (ALT) $> 5 \times$ upper limit of normal (ULN);<br>b. Total bilirubin $> 1.5 \times$ ULN, unless Gilbert’s syndrome is present in which case total bilirubin $> 2 \times$ ULN.<br>7. Known or persistent use, misuse, or dependency of medication, drugs, or alcohol.<br>8. Current or planned pregnancy or women who are breastfeeding.<br>9. Patients with severe vision or hearing impairment (that is not corrected by glasses or hearing aids) that, at the investigator’s discretion, interferes with their ability to perform study assessments.<br>10. Patients who have been diagnosed with arthritis or other musculoskeletal disorders affecting joints, muscles, ligaments, and/or nerves that by themselves affects patient’s mobility and, at the investigator’s discretion, interferes with their ability to perform study assessments. |
| 3. Females of childbearing potential, defined as a premenopausal female capable of becoming pregnant, will be included if they are either sexually inactive (sexually abstinent<sup>1</sup> for 14 days prior to the first dose and confirm to continue through 28 days after the last dose) or using one of the following highly effective contraceptives (i.e. results in <1% failure rate when used consistently and correctly) 14 days prior to the first dose continuing through 28 days after the last dose:<br>a) intrauterine device (IUD);<br>b) surgical sterilization of the partner (vasectomy for 6 months minimum);<br>c) combined (estrogen or progestogen containing) hormonal contraception associated with the inhibition of ovulation (either oral, intravaginal, or transdermal);<br>d) progestogen only hormonal contraception associated with the inhibition of ovulation (either oral, injectable, or implantable);<br>e) intrauterine hormone releasing system (IUS);<br>f) bilateral tubal occlusion.<br>4. Females of non-childbearing potential must have undergone one of the following sterilization procedures at least 6 months prior to the first dose:<br>a) hysteroscopic sterilization;<br>b) bilateral tubal ligation or bilateral salpingectomy;<br>c) hysterectomy;<br>d) bilateral oophorectomy;<br>**OR** be postmenopausal with amenorrhea for at least 1 year prior to the first dose and follicle stimulating hormone (FSH) serum levels consistent with postmenopausal status. FSH analysis for postmenopausal women will be done at screening. FSH levels should be in the postmenopausal range as determined by the central laboratory.<br>5. Non-vasectomized male patient agrees to use a condom with spermicide or abstain from sexual intercourse during the study until 90 days beyond the last dose of study medication and the female partner agrees to comply with inclusion criteria 3 or 4. For a vasectomized male who has had his vasectomy 6 months or more prior to study start, it is required that they use a condom during sexual intercourse. A male who has been vasectomized less than 6 months prior to study start must follow the same restrictions as a non-vasectomized male. | 11. Patients unwilling and/or not able to undergo a 42-day washout period from any of the following prohibited medication prior to **Visit 1** (Baseline 1) and remain without prohibited medication through **Visit 6**.<br>a) Aminopyridines (including sustained-release form);<br>b) N-Acetyl-DL-Leucine (e.g. Tanganil<sup>®</sup>) ;<br>c) N-Acetyl-L-Leucine (prohibited if not provided as IMP);<br>d) Riluzole;<br>e) Gabapentin;<br>f) Varenicline;<br>g) Chlorzoxazone;<br>h) Sulfasalazine;<br>i) Rosuvastatin. |
| 6. If male, patient agrees not to donate sperm from the first dose until 90 days after their last dose.<br>7. Patients must fall within:<br>a) A SARA score of $5 \leq X \leq 33$ points (out of 40)<br>**AND**<br>b) Either:<br>i. Within the 2-7 range (0-8 range) of the Gait subtest of the SARA scale<br>**OR**<br>ii. Be able to perform the 9-Hole Peg Test with Dominant Hand (9HPT-D) (SCAFI subtest) in $20 \leq X \leq 150$ seconds.<br>8. Weight $\geq 15$ kg at screening.<br>9. Patients are willing to disclose their existing medications/therapies for (the symptoms) of NPC, including those on the prohibited medication list. Non-prohibited medications/therapies (e.g. miglustat, concomitant speech therapy, and physiotherapy) are permitted provided:<br>a) The Investigator does not believe the medication/therapy will interfere with the study protocol/results<br>b) Patients have been on a stable dose/duration and type of therapy for at least 42 days before **Visit 1** (Baseline 1)<br>c) Patients are willing to maintain a stable dose/do not change their therapy throughout the duration of the study.<br>10. An understanding of the implications of study participation, provided in the written patient information and informed consent by patients or their legal representative/parent, and demonstrates a willingness to comply with instructions and attend required study visits (for children this criterion will also be assessed in parents or appointed guardians). |  |

<sup>1</sup> Sexual abstinence is considered a highly effective method only if defined as refraining from heterosexual intercourse during the entire period of risk associated with the study treatments. In this trial abstinence is only acceptable if in line with the patient's preferred and usual lifestyle.
Periodic abstinence (calendar, symptothermal, post-ovulation methods), withdrawal (coitus interruptus), spermicides only, and lactational amenorrhoea method (LAM) are not acceptable methods of contraception. As well, female condom and male condom should not be used together.

Approval was obtained by the applicable responsible central research ethics committees / institutional review boards for each center. Written informed consent was obtained for all study participants by the subject or, if applicable, their parent or legal representative.

### Outcome Measures

#### Primary endpoint

The primary endpoint was the novel, functionally relevant Clinical Impression of Change in Severity (CI-CS). The CI-CS assessment was performed by independent, blinded raters who compared videos of each patient performing a “primary anchor test” (either the 8 Meter Walk Test (8MWT) or 9 Hole Peg Test – Dominant Hand (9HPT-D)) at baseline, treatment, and washout study visits. Blinded to the timepoint of each video, the raters made an objective comparison scored on a 7-point Likert scale of the change in the severity of the patient’s neurological signs and symptoms from Video 1 to Video 2. Details of CI-CS administration and assessment have been previously published (Fields et al., 2020).

Each patient’s primary anchor test (8MWT or 9HPT-D) was selected by the Principal Investigator at Visit 1 based on the patient’s unique clinical symptoms to better ensure the clinically relevancy of the primary outcome assessment. The anchor tests were filmed in a standardized way at each visit and uploaded for centralized review. A pool of three board certified neurologists performed the central video analysis. Two “primary raters” were responsible for the initial comparison of the video pairs. For cases where the two initial reviewers differed in their assessment of the primary CI-CS score by more than one (1) point, the third rater acted as an adjudicator.

The raters assessed: “Compared to the first video, how has the severity of the patient’s performance on the 8MWT or 9HPT-D changed (improved or worsened) as observed in the second video?” The CI-CS assessment was based on a 7-point Likert scale ranging from +3 (significantly improved) to −3 (significantly worse).

#### Secondary Outcomes

Secondary assessments were assessed and analyzed between two time points: (i) baseline (day 0) to the end of treatment (approximately day 42); the end of treatment (approximately day 42) to the end of post-treatment washout (day 84).

To evaluate the overall neurologic status in NPC disease, the modified Disability Rating Scale (mDRS) was applied, consisting of 6 subdomains (ambulation, manipulation, Seizures, Language, Swallowing, Ocular movements) (Iturriaga et al., 2006). Cerebellar function evaluations were administered, including: (1) the Scale for the Assessment and Rating of Ataxia (SARA), an 8-item clinical rating scale (gait, stance, sitting, speech, fine-motor function, and coordination; range 0– 40, where 0 is the best neurologic status and 40 the worst) (Schmitz-Hübsch et al., 2006); and (2) the Spinocerebellar Ataxia Functional Index (SCAFI), comprising 8-m walking time performed by having patients walk twice, as quickly as possible, from one line to another excluding turning, the 9-Hole Peg Test (9HPT) with the dominant and non-dominant hand, and the number of “PATA” repetitions over 10 seconds (PATA) (Schmitz-Hübsch et al., 2008).

Subjective impairment and quality of life were evaluated by using the Clinical Global Impression of Severity and Improvement Scale (completed by the Investigator, Caregiver, and Subject) (Quinn et al., 2008), the EuroQol (EQ) 5Q-5D-5L/Y questionnaire and the visual analog scale (VAS) (EuroQol Group, 2017).

### Statistical Analysis

The primary endpoint was defined as the numerical difference (delta, Δ) of the CI-CS value for the treatment period (Visit 2 versus Visit 4) minus the CI-CS value for the washout period (Visit 4 versus Visit 6). Values of this endpoint would be positive for example if the patient improved or remained stable through the treatment period but then remained stable or worsened respectively during the washout period, while negative values would result if a patient worsened or remained stable during the treatment period but then remained stable or improved respectively during the washout period. The primary analysis was performed according to the modified intention-to-treat (mITT) principle, used to estimate the treatment effect regardless of discontinuation and provides a perspective of the treatment effect across the entire population. The mITT analysis set was defined as all patients who receive at least one dose of the study drug and with one video recording during the baseline period (Visit 1 or 2, or both) and treatment period (Visit 3 or Visit 4, or both). The mITT analysis utilized a last observation carried forward approach for the primary CI-CS endpoint which implies that the CI-CS value for Visit 4 to Visit 6 is assigned the value 0 (stable) if both videos from the washout period (Visit 5 and Visit 6) are not available. The analysis of the primary endpoint was based on a single sample one-sided Wilcoxon comparing the mean of the CI-CS differences with zero. The null hypothesis is that the mean is ≤0, with the alternative hypothesis that this mean is >0 and the test will be conducted at the one-sided 5% significance level. Non-parametric 90% confidence intervals were constructed using the Hodges-Lehmann method (Hodges and Lehmann, 1963). Secondary endpoints, which included the separate CI-CS scores during treatment and washout and separate SARA scores and the CGI-C during treatment and washout were evaluated either statistically based on a single sample t-test or a single sample Wilcoxon Signed Rank test or descriptively, as pre-defined in the Statistical Analysis Plan (SAP). There was no formal hierarchical structure defined for the secondary endpoints and results on these endpoints should therefore be viewed as exploratory. For each of the primary and secondary endpoints there will be separate analyses within key subgroups pre-defined in the SAP. These subgroups are listed in **Figure 2**.

**Figure 2.**
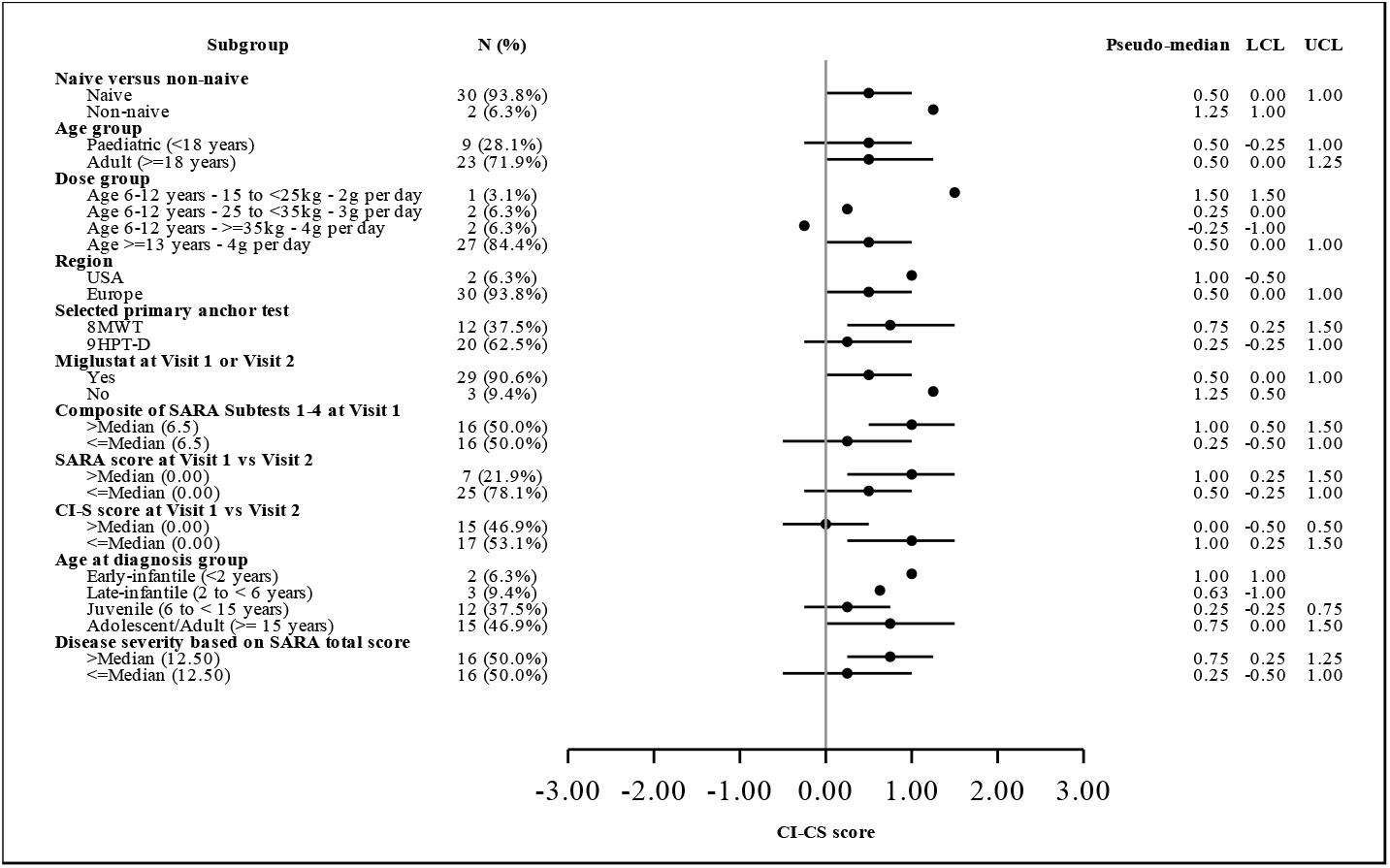
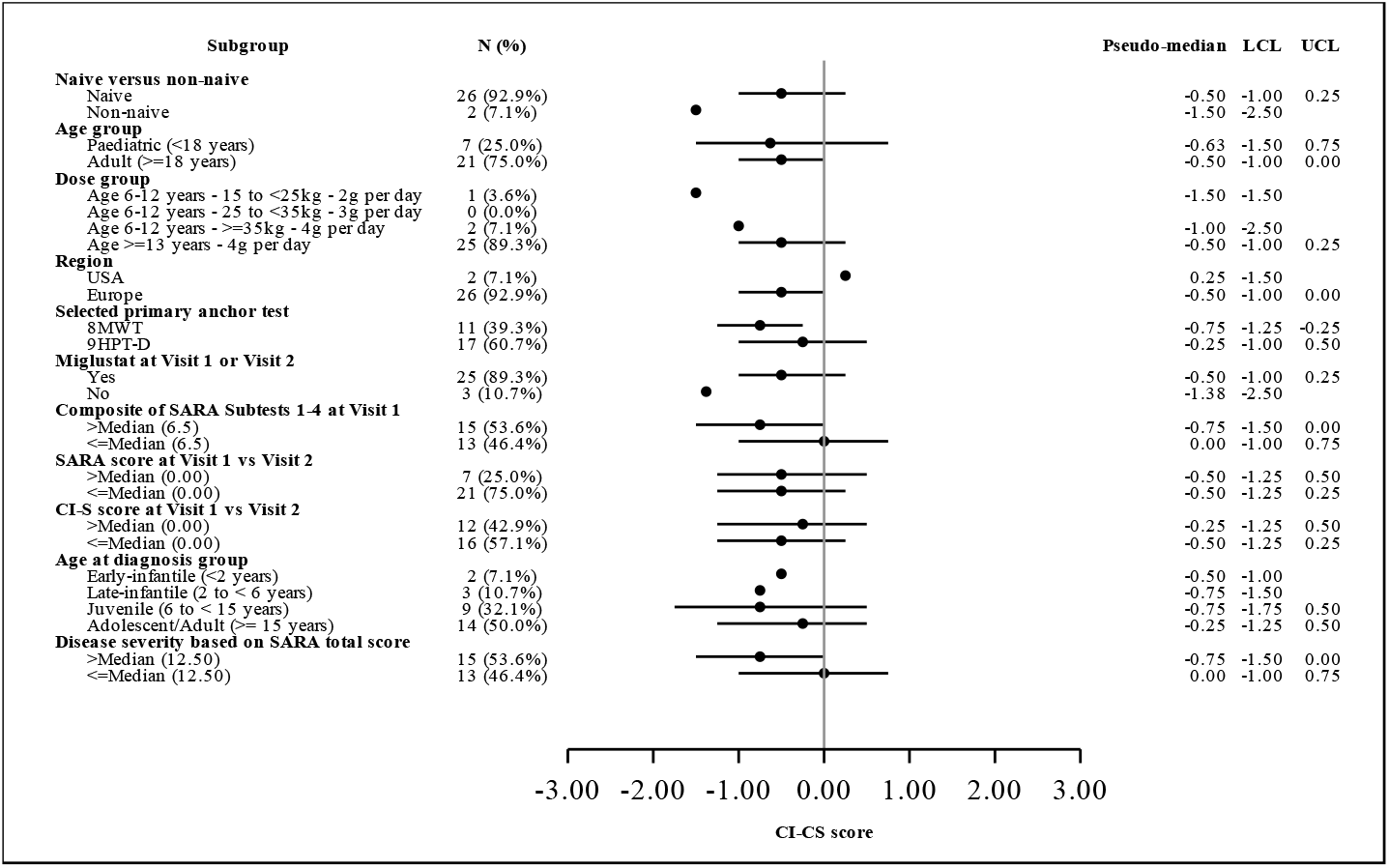
Forest plot for CI-CS scores for pre-defined subgroup analysis, based on the mITT population. The dots represent the pseudo-medians or Hodges-Lehmann estimators, the horizontal lines represent the 90% confidence intervals. LCL = Lower confidence limit, UCL = Upper confidence limit. For some subgroups the number of patients was too small to calculate the LCL and/or UCL, in that case the result is presented as missing and no line presenting the confidence interval is drawn. No last observation carried forward (LOCF) approach was used for this figure. Only values from patients with reported data are included. **A)**. CI-CS scores visit 4 (end of treatment) vs. Visit 2 (baseline). **B)**. CI-CS scores visit 6 (end of washout) vs. Visit 4 (end of treatment).

## RESULTS

### Characteristics of the Patients

Thirty-four participants were screened, and thirty-three subjects were enrolled per protocol. The demographic and baseline clinical characteristics of the patients are presented in **Table 2**.

**Table 2.** Demographics and Baseline Characteristics

|  |  |  |
| --- | --- | --- |
| **Age (years)** | Mean (SD) | 28.8 (15.1) |
|  | Median | 29 |
|  | Range | 7.0 – 64.0 |
| **Ethnicity, n (%)** | Asian | 1 (3.0%) |
|  | White | 30 (90.9%) |
|  | Other | 2 (6.1%) |
| **Gender, n (%)** | Male | 24 (72.7%) |
|  | Female | 9 (27.3%) |
| **Age Group, n (%)** | Paediatric (<18 years) | 10 (30.3%) |
|  | Adult (≥18 years) | 23 (69.7%) |
| **Dose, n (%)** | Age 6-12 years - 15 to <25kg - 2g per day | 1 (3.0%) |
|  | Age 6-12 years - 25 to <35kg - 3g per day | 2 (6.1%) |
|  | Age 6-12 years - ≥35kg - 4g per day | 2 (6.1%) |
|  | Age ≥13 years - 4g per day | 28 (84.8%) |
| **Geographic Location, n (%)** | USA | 2 (6.1%) |
|  | Europe | 31 (93.9%) |
| **Miglustat at baseline, n (%)** | Yes | 30 (90.9%) |
|  | No | 3 (9.1%) |
| **Selected Primary Anchor Test, n (%)** | 8 Meter Walk Test (8MWT) | 12 (36.4%) |
|  | 9 Hole Peg Test – Dominant Hand (9HPT-D) | 21 (63.6%) |

Thirty-two patients qualified for the primary mITT analysis set (96.9%), which included all patients dosed who had at least one treatment visit (Visit 3 or Visit 4). One patient was withdrawn after Visit 2 due to an unrelated adverse event (epileptic seizures). One patient was withdrawn between Visit 3 and Visit 4 following an unrelated serious adverse event (fall, resulting in head injury/trauma and two broken ribs). The median duration of IB1001 treatment was 43 days (range: 35-133), with a mean duration of exposure of 51.7 days. The COVID-19 pandemic impacted the duration of treatment /post-treatment washout for select patients, and patients were dosed / on washout until it was safe and feasible to conduct an on-site end of treatment / washout visit in adherence with all COVID-19 local regulations. These deviations due to COVID-19 are reflected by the limited size of the per protocol population (n = 18). Thirty-one patients (93.9%) completed the Parent Study (Visit 6).

### Primary Outcome

The CI-CS primary endpoint of the study reached statistical significance with *p*=0.029 with mean value=0.86 (SD=2.52, median=1.0) and Hodges-Lehmann 90% confidence interval (CI) (0.25, 1.75). There were no missing values for CI-CS for the treatment period although there were four missing values for CI-CS during the washout period.

The CI-CS component for the treatment period (Visit 4 versus Visit 2) had a mean value of 0.48 (SD=1.34 median=1.0), showing on average an improvement in the patients’ condition over that period, while the CI-CS component for the washout period (Visit 6 versus Visit 4) had a mean value of −0.38 (SD=1.45, median= −0.25) showing on average a worsening. There was no difference observed between the CI-CS comparing the baseline and washout visits (Visit 6 versus Visit 2): mean value 0.063 (SD=1.32, median =0, n=30).

### Secondary Outcomes

The mean total SARA score at baseline (Visit 2) was 14.47 (SD=7.14, median=12.74, n=32) and encompassed a full range of disease-severity (min=5, max=33). The mean change in score during the treatment period (Visit 2 to Visit 4) was −1.19 (SD=2.02, median = −1.00, n=31) with 90% CI (−1.75, −0.50) *p*=0.001, demonstrating an improvement on cerebellar signs and neurological symptoms. The mean change in score during the washout period (Visit 4 to Visit 6) was 1.45 (SD=2.56, median 0.75, n=28) with 90% CI (0.50, 2.00), *p*=0.002, showing a deterioration. There was no difference observed between the baseline visit to the washout visit (Visit 2 to Visit 6); the mean change in score was 0.25 (SD=2.60, median=0, n=28) with 90% CI (1.1, −0.59) *p*=0.308.

The CGI-Change is presented around a reference value of 0=no change, so that for example - 1=minimally worse and +1=minimally improved. The investigator, caregiver, and patient CGI-C were consistent and showed an average improvement during the treatment period and deterioration during the washout period. For the investigator’s CGI-C, the mean change from Visit 2 to Visit 4 was 0.6 (SD=0.9, median=0, n=31) with 90% CI (0.5, 1.0) *p*<0.001 while the mean change from Visit 4 to Visit 6 was −0.5 (SD=0.9, median = 0, n=28) with 90% CI (−1.0, 0) *p*=0.006. For the caregiver’s CGI-C, the mean change from Visit 2 to Visit 4 was 0.6 (SD=1.0, median=0, n=30) with 90% CI (1.0, 0.0) *p*=0.005, while the mean change from Visit 4 to Visit 6 was −0.4 (SD=1.1, median = 0, n=27) with 90% CI −1.0, 0.0) *p*=0.038. For the patient’s CGI-C, the mean change from Visit 2 to Visit 4 was 0.7 (SD=1.0, median=0.5, n=25) with 90% CI (0.5, 1.0) *p*=0.003, while the mean change from Visit 4 to Visit 6 was −0.4 (SD=0.9, median = 0, n=24) with 90% CI (−0.5, 0.0) *p*=0.034.

The mean total SCAFI score at baseline (Visit 2) was −0.3011 (SD=1.0405, median=-0.1362, n=32). The score during the treatment period showed a trend towards improvement, with a mean change (Visit 2 to Visit 4) of 0.0995 (SD=0.3058, median = 0.0890, n=30) with 90% CI (−0.0123, 0.1777) *p*=0.084. The mean change in score during the washout period (Visit 4 to Visit 6) was 0.0076 (SD=0.3584, median −0.0621, n=26) with 90% CI (−0.1269, 0.1031) *p*=0.315. There was a trend for improvement on the 8MWT (mean −1.093, 90% CI −1.53, 0.05, *p*=0.065) and PATA test (mean 0.95, 90% CI 0.0, 1.75, *p*=0.076) on medication. As reported, the quantifiable time-based SCAFI assessments do not best capture clinically meaningful changes in functioning or quality of life; hence, the CI-CS assessment was developed (Koch et al., 2013).

The mean mDRS at baseline was 0.467 (SD=0.155, median=0.458, n=32). Change from baseline through to Visit 4 showed a small improvement in terms of disability on average with a mean change of −0.012 (SD=0.050, median=0, n=31), 90% CI (−0.13, 0.08) *p*=0.121 with a mean change from Visit 4 to Visit 6 (washout) of 0.016 (SD-0.051, median=0, n=28), 90% CI (0.0, 0.042), p=0.056 on average showing a slight worsening in terms of disability.

A Forest plot (**Figure 2**) is included to display results for the primary endpoint in the pre-defined subgroups showing Hodges-Lehman median estimates and corresponding 90% confidence intervals calculated where possible. There is some variation as would be expected given the small sample sizes but there is no evidence that would suggest a differential treatment benefit across the population as a whole. Instead, all demographics demonstrated the consistent trends of improving during treatment and deterioration during the washout.

### Safety parameters

Results of serum and urine tests and ECG recordings were normal or rated as clinically non-significant. Adherence was high as shown by treatment compliance and the regular urine analyses for prohibited medication.

### Adverse Events

A treatment emergent adverse event (TEAE) was any adverse event (AE) that appeared or worsened after study treatment began (i.e. in the treatment or washout period). The distribution of TEAEs is shown in **Table 3**. There were no AEs with an incidence >10%. Seven related AEs were reported for 4 patients, including: flatulence, diarrhoea, increased feeling of hunger, rash (twice for 1 patient), and aggressive behaviour accompanied by restlessness. The events were transient and manageable. No serious adverse reactions were reported. No deaths occurred during the study.

**Table 3.** Distribution of Treatment-emergent adverse events (TEAEs) based on the Safety Analysis Set (SAF)

| System Organ Class<br>Preferred Term | Total (N=33) |  |
| --- | --- | --- |
|  | n (%) | m |
| Any treatment emergent adverse event | 24 (72.7%) | 57 |
| Infections and infestations | 9 (27.3%) | 10 |
| Gastroenteritis | 2 (6.1%) | 2 |
| Lower respiratory tract infection | 2 (6.1%) | 2 |
| Nasopharyngitis | 1 (3.0%) | 1 |
| Rhinitis | 3 (9.1%) | 3 |
| Upper respiratory tract infection | 2 (6.1%) | 2 |
| Nervous system disorders | 8 (24.2%) | 10 |
| Balance disorder | 1 (3.0%) | 1 |
| Coordination abnormal | 1 (3.0%) | 1 |
| Dementia | 1 (3.0%) | 1 |
| Drooling | 1 (3.0%) | 1 |
| Dropped head syndrome | 1 (3.0%) | 1 |
| Headache | 1 (3.0%) | 1 |
| Petit mal epilepsy | 1 (3.0%) | 1 |
| Seizure | 3 (9.1%) | 3 |
| Injury, poisoning and procedural complications | 5 (15.2%) | 9 |
| Contusion | 1 (3.0%) | 1 |
| Fall | 3 (9.1%) | 5 |
| Head injury | 1 (3.0%) | 1 |
| Rib fracture | 1 (3.0%) | 1 |
| Traumatic haematoma | 1 (3.0%) | 1 |
| Gastrointestinal disorders | 5 (15.2%) | 8 |
| Anal incontinence | 1 (3.0%) | 1 |
| Diarrhoea | 3 (9.1%) | 5 |
| Dysphagia | 1 (3.0%) | 1 |
| Flatulence | 1 (3.0%) | 1 |
| Respiratory, thoracic and mediastinal disorders | 5 (15.2%) | 5 |
| Cough | 1 (3.0%) | 1 |
| Epistaxis | 2 (6.1%) | 2 |
| Obstructive airways disorder | 1 (3.0%) | 1 |
| Productive cough | 1 (3.0%) | 1 |
| Skin and subcutaneous tissue disorders | 3 (9.1%) | 4 |
| Rash | 2 (6.1%) | 2 |
| Rash pruritic | 2 (6.1%) | 2 |
| General disorders and administration site conditions | 2 (6.1%) | 2 |
| Asthenia | 1 (3.0%) | 1 |
| Fatigue | 1 (3.0%) | 1 |
| Investigations | 2 (6.1%) | 2 |
| Blood alkaline phosphatase increased | 1 (3.0%) | 1 |
| Blood pressure increased | 1 (3.0%) | 1 |
| Metabolism and nutrition disorders | 2 (6.1%) | 2 |
| Dehydration | 1 (3.0%) | 1 |
| Increased appetite | 1 (3.0%) | 1 |
| Musculoskeletal and connective tissue disorders | 2 (6.1%) | 2 |
| Arthralgia | 1 (3.0%) | 1 |
| Neck pain | 1 (3.0%) | 1 |
| Psychiatric disorders | 1 (3.0%) | 2 |
| Aggression | 1 (3.0%) | 1 |
| Restlessness | 1 (3.0%) | 1 |
| Reproductive system and breast disorders | 1 (3.0%) | 1 |
| Ejaculation failure | 1 (3.0%) | 1 |

## DISCUSSION

Here we report the results of a Phase II clinical trial investigating the safety and efficacy of the modified amino acid NALL for NPC disease in patients aged 6 years or older. The major findings of the IB1001-201 trial are: first, NALL improved symptoms, including gait and stance, upper extremity and fine motor function, which worsened during the post-treatment wash-out. Second, consistent with its pharmacological action, NALL improved cerebellar signs after 6 weeks. Third, improvement of neurological status was observed across all demographics of patients (age, gender, age of onset, disease severity, etc.) establishing a rational for IB1001 to be used as a treatment for all NPC patients. Fourth, the low frequency (7 related AEs in 4 patients of 32 participants) and transient, mild nature of these AEs inform a favorable benefit-risk profile.

Methodologically, to elaborate a novel clinical endpoint, the CI-CS based on videos of the 8MWT or the 9HPT-D was used which was rated (0±3) by independent, blinded neurologists. In addition to reducing detection and performance bias for the primary endpoint, the blinded CI-CS served as a metric of clinical importance that could not be obtained from the traditional timed assessments. Instead, raters evaluated clinically meaningful changes in patient’s neurological manifestations which correlate to their level of functioning and quality of life, such as the accuracy and fluidity of movements, spasticity, ataxia, and dystonia for the 9HPT-D, and changes in gait patterns such as balance and postural stability, variability, asymmetry, ataxia, and support for the 8MWT.

Of note, 93.8% of patients were on the standard of care agent Miglustat. The administration of NALL therefore showed significant effects beyond Miglustat for the signs and symptoms of NPC. This is in line with the previous observational studies that indicated the additive effect of NALL with Miglustat (Bremova et al., 2015; Cortina-Borja et al., 2018), and studies demonstrating synergistic effects of NALL and Miglustat (Kaya et al., 2020a).

NALL’s positive treatment effects directly correlate to its pharmacological action: Animal studies in an *Npc1*^-/-^ mouse model show NALL significantly reduces ataxia when treatment is initiated either symptomatically (from 8-weeks of age) or pre-symptomatically (from 3-weeks of age) (Kaya et al., 2020a). These *in vivo* studies further show NALL can restore neuronal function and protect against/delay disease progression in multiple neurological brain circuits. Altered glucose and antioxidant metabolism and reduction of cerebellar inflammation have been implicated as potential mechanisms of action (Kaya et al., 2020a; Sarkar et al., 2019). The clinical manifestations of this, as demonstrated in the IB1001-201 clinical trial, were visualized and captured in terms of improvements or stabilization in very different processes such as ambulation, fine motor skills, speech, and cognition.

The IB1001-201 parent study has several limitations. First, it was not placebo-controlled. To prevent potential patient expectation or investigator bias, all aspects of the administration and videorecording of the CI-CS anchor tests were standardized to ensure the quality of videos assessed by the blinded raters. Prior to any patient visit, all sites were trained on these detailed protocols, including precise verbal instructions, encouragement, break times between test trials, and instructions on which trial to video. Given the majority of NPC patients enrolled in the IB1001-201 clinical trial featured severe physical impairments with regard to both their fine motor skills as well as balance and gait, and mild to significant levels of cognitive impairment, the potential for a placebo-effect which significantly altered neurological signs and symptoms–the basis of the CI-CS assessment – was therefore considered minimal, ensuring the interpretability of the blinded-raters’ CI-CS assessments.

Second, the novel CI-CS primary endpoint has not been previously used or yet validated. It was implemented, however, due to the methodological limitations of applying pre-existing ataxia scales in heterogenous diseases, where the scales may not be too broad and therefore not sensitive to capture small but meaningful functional changes (Fields et al., 2020; Perez-Lloret et al., 2020). The validated NPC Clinical Severity Scale is only validated to measure disease progression after a minimum of 1 year; thus, it is not sensitive enough a measure to capture change in after 6-weeks (Yanjanin et al., 2010). Accordingly, the CI-CS was developed to be a more clinically relevant endpoint capable of detecting clinically meaningful treatment effects. The CI-CS was demonstrated to be a reliable instrument, with very high inter-rater consistency (0.80 correlation).

Third, this study was limited to an investigation of the symptomatic effect of NALL treatment; data from the ongoing Extension Phase will provide further insights into effects on disease progression and long-term safety.

Consistent with previous non-clinical and observational clinical studies, IB1001 demonstrated a significant and clinically meaningful improvement of symptoms, functioning, and quality of life in pediatric and adult NPC patients. IB1001 was safe and well-tolerated, contributing to a favorable benefit-risk profile for the treatment of this serious, debilitating disease.

## Supporting information

CONSORT Checklist

## Data Availability

The datasets generated during and/or analysed during the current study are available from IntraBio Ltd. Due to confidentiality agreements with research collaborators, the data are not publicly available but are available from the corresponding author and with permission from IntraBio Ltd on reasonable request.

## Author Contributions

TF, JGB, PG, AH Hahn, AH Hassan, AH Hennig, SJ, MK, KM, UR, RS, SS were responsible for the recruitment and examination of subjects/ acquisition of subject data. TBE and JC were responsible for the assessment of the primary data. TBE and SAS drafted the manuscript. TBE, JC, TF, JGB, PG, AH Hahn, AH Hassan, AH Hennig, SJ, MK, KM, UR, RS, SS revised the manuscript for content and provided final approval of the manuscript.

## Acknowledgements

We thank each study team and co-investigators for their participation in the trial. We thank the multinational Patient Organizations representing the NPC community and referring physicians. Finally, we thank all of the patients and their families who participated in this study.

## REFERNCES

Bremova, T., Malinová, V., Amraoui, Y., Mengel, E., Reinke, J., Kolníková, M., and Strupp, M. (2015). Acetyl-dl-leucine in Niemann-Pick type C: A case series. Neurology 85, 1368–1375.

Churchill, G.C., Strupp, M., Galione, A., and Platt, F.M. (2020). Unexpected differences in the pharmacokinetics of N-acetyl-DL-leucine enantiomers after oral dosing and their clinical relevance. PLoS One 15, e0229585.

Cortina-Borja, M., Te Vruchte, D., Mengel, E., Amraoui, Y., Imrie, J., Jones, S.A.I Dali, C., Fineran, P., Kirkegaard, T., Runz, H., et al. (2018). Annual severity increment score as a tool for stratifying patients with Niemann-Pick disease type C and for recruitment to clinical trials. Orphanet J Rare Dis 13, 143.

EuroQol Group (2017). EQ-5D Instruments | About EQ-5D. 2017.

Fields, T., Patterson, M., Bremova, T., Belcher, G., Billington, I., Churchill, G.C., Davis, W., Evans, W., Flint, S., Galione, A., et al. (2020). A master protocol to investigate a novel therapy acetyl-L-leucine for three ultra-rare neurodegenerative diseases: Niemann-Pick type C, the GM2 Gangliosidoses, and Ataxia Telangiectasia. MedRxiv 2020.03.11.20034256.

Geberhiwot, T., Moro, A., Dardis, A., Ramaswami, U., Sirrs, S., Marfa, M.P., Vanier, M.T., Walterfang, M., Bolton, S., Dawson, C., et al. (2018). Consensus clinical management guidelines for Niemann-Pick disease type C. Orphanet J Rare Dis 13, 50.

Günther, L., Beck, R., Xiong, G., Potschka, H., Jahn, K., Bartenstein, P., Brandt, T., Dutia, M., Dieterich, M., Strupp, M., et al. (2015). N-acetyl-L-leucine accelerates vestibular compensation after unilateral labyrinthectomy by action in the cerebellum and thalamus. PLoS ONE 10, e0120891.

Hodges, J.L., and Lehmann, E.L. (1963). Estimates of Location Based on Rank Tests. The Annals of Mathematical Statistics 34, 598–611.

Iturriaga, C., Pineda, M., Fernández-Valero, E.M., Vanier, M.T., and Coll, M.J. (2006). Niemann-Pick C disease in Spain: clinical spectrum and development of a disability scale. J. Neurol. Sci. 249, 1–6.

Kaya, E., Smith, D.A., Smith, C., Morris, L., Bremova-Ertl, T., Cortina-Borja, M., Fineran, P., Morten, K.J., Poulton, J., Boland, B., et al. (2020a). Acetyl-leucine slows disease progression in lysosomal storage disorders. Brain Commun fcaa148, https://doi.org/10.1093/braincomms/fcaa148.

Kaya, E., Smith, D.A., Smith, C., Boland, B., Strupp, M., and Platt, F.M. (2020b). Beneficial effects of acetyl-DL-leucine (ADLL) in a mouse model of Sandhoff disease. J Clin Med 9, 1050.

Koch, M.W., Cutter, G., Stys, P.K., Yong, V.W., and Metz, L.M. (2013). Treatment trials in progressive MS--current challenges and future directions. Nat Rev Neurol 9, 496–503.

Patterson, M.C., Clayton, P., Gissen, P., Anheim, M., Bauer, P., Bonnot, O., Dardis, A., Dionisi-Vici, C., Klünemann, H.-H., Latour, P., et al. (2017). Recommendations for the detection and diagnosis of Niemann-Pick disease type C: An update. Neurol Clin Pract 7, 499–511.

Perez-Lloret, S., van de Warrenburg, B., Rossi, M., Rodríguez-Blázquez, C., Zesiewicz, T., Saute, J.A.M., Durr, A., Nishizawa, M., Martinez-Martin, P., Stebbins, G.T., et al. (2020). Assessment of Ataxia Rating Scales and Cerebellar Functional Tests: Critique and Recommendations. Mov Disord.

Quinn, T.J., Dawson, J., Walters, M.R., and Lees, K.R. (2008). Variability in modified Rankin scoring across a large cohort of international observers. Stroke 39, 2975–2979.

Sarkar, C., Hegdekar, N., and Lipinski, M.M. (2019). N-acetyl-L-leucine treatment attenuates neuronal cell death and suppresses neuroinflammation after traumatic brain injury in mice. BioRxiv 759894.

Schmitz-Hübsch, T., du Montcel, S.T., Baliko, L., Berciano, J., Boesch, S., Depondt, C., Giunti, P., Globas, C., Infante, J., Kang, J.-S., et al. (2006). Scale for the assessment and rating of ataxia: development of a new clinical scale. Neurology 66, 1717–1720.

Schmitz-Hübsch, T., Giunti, P., Stephenson, D.A., Globas, C., Baliko, L., Saccà, F., Mariotti, C., Rakowicz, M., Szymanski, S., Infante, J., et al. (2008). SCA Functional Index: a useful compound performance measure for spinocerebellar ataxia. Neurology 71, 486–492.

Tighilet, B., Leonard, J., Bernard-Demanze, L., and Lacour, M. (2015). Comparative analysis of pharmacological treatments with N-acetyl-DL-leucine (Tanganil) and its two isomers (N-acetyl-L-leucine and N-acetyl-D-leucine) on vestibular compensation: Behavioral investigation in the cat. Eur. J. Pharmacol. 769, 342–349.

Vanier, M.T., and Millat, G. (2003). Niemann-Pick disease type C. Clin. Genet. 64, 269–281.

Yanjanin, N.M., Vélez, J.I., Gropman, A., King, K., Bianconi, S.E., Conley, S.K., Brewer, C.C., Solomon, B., Pavan, W.J., Arcos-Burgos, M., et al. (2010). Linear clinical progression, independent of age of onset, in Niemann-Pick disease, type C. Am J Med Genet B Neuropsychiatr Genet 153B, 132–140.

