## Supplementary material for "Acetyl-L-leucine for Niemann-Pick type C – a multi-national, rater-blinded phase II trial": CONSORT Checklist

### CONSORT 2010 checklist of information to include when reporting a randomised trial*

| Section/Topic | Item No | Checklist item | Reported on page No |
|---|---|---|---|
| **Title and abstract** | | | |
| | 1a | Identification as a randomised trial in the title | No randomization/allocation of subjects to different intervention group occurred. The only randomization which occurred for the IB1001 clinical trials was the randomization of subject videos for central, independent raters primary CI-CS assessment. Therefore, “randomization” was not included in the trial, as to not be misleading about subject allocation to different intervention arms. |
| | 1b | Structured summary of trial design, methods, results, and conclusions (for specific guidance see CONSORT for abstracts) | 3 |
| **Introduction** | | | |
| Background and objectives | 2a | Scientific background and explanation of rationale | 4-5 |
| | 2b | Specific objectives or hypotheses | 5 |
| **Methods** | | | |
| Trial design | 3a | Description of trial design (such as parallel, factorial) including allocation ratio | 5 |
| | 3b | Important changes to methods after trial commencement (such as eligibility criteria), with reasons | 5 |
| Participants | 4a | Eligibility criteria for participants | 5, 14,15, 16 |
| | 4b | Settings and locations where the data were collected | 5 |
| Interventions | 5 | The interventions for each group with sufficient details to allow replication, including how and when they were actually administered | 5 |
| Outcomes | 6a | Completely defined pre-specified primary and secondary outcome measures, including how and when they were assessed | 6,7 |
| | 6b | Any changes to trial outcomes after the trial | No changes to the trial outcomes after the trial commenced |

| Section/Topic | Item No | Checklist item | Reported on page No |
|---|---|---|---|
| | | commenced, with reasons | |
| Sample size | 7a | How sample size was determined | Separate document – Statistical Analysis Plan |
| | 7b | When applicable, explanation of any interim analyses and stopping guidelines | No interim analyses occurred |
| Randomisation: | | | |
| Sequence generation | 8a | Method used to generate the random allocation sequence | No randomization/allocation of subjects to different intervention group occurred. The only randomization which occurred for the IB1001 clinical trials was the randomization of subject videos for central, independent raters primary CI-CS assessment. This randomization/release to raters was performed by a third-party vendor and detailed in the Separate Document -Video Review Charter. |
| | 8b | Type of randomisation; details of any restriction (such as blocking and block size) | The following information is available in external documents (Video Review Charter): The primary Clinical Impression of Change in Severity (CI-CS) assessment was based on videos of subjects recorded at each study visit. These videos were uploaded directly from each study site to a third-party vendor, Medpace Core Laboratories (MCL). MCL was responsible for obtaining subject videos, performing quality control, and, once all applicable videos had been obtained for a subject, assigning the subject videos to randomized sequences for the pairs of videos for the CI-CS assessment.<br><br>In advance of any video review, MCL utilizes a number generator via *RANDOM.ORG* to generate a random number (1-6) for each of the participants. The generated number for each patient (1-6) corresponds to the CI-CS video review order assignments initially outlined in the video review charter (below)<br><br>(table below)<br><br>Once a subject's videos are randomized according to the generated order, these sets |

| | Video A | Video B | Video C |
|---|---|---|---|
| Order 1 | 2 | 4 | 6 |
| Order 2 | 2 | 6 | 4 |
| Order 3 | 4 | 2 | 6 |
| Order 4 | 4 | 6 | 2 |
| Order 5 | 6 | 2 | 4 |
| Order 6 | 6 | 4 | 2 |

| | | | |
|---|---|---|---|
| | | | were released via Clintrak Imaging System Portal for review by independent raters. The independent raters based their primary CI-CS on the review of these randomized video /pairs.<br><br>The procedures for acquiring subject videos, submitting the videos to MCL, and central video review are described in the separate documents (Video Acquisition Manual, Video Submission Manual, Video Review Charter). |
| Allocation concealment mechanism | 9 | Mechanism used to implement the random allocation sequence (such as sequentially numbered containers), describing any steps taken to conceal the sequence until interventions were assigned | The video randomization sequences for the CI-CS assessment was pre-defined by MCL anda biostatistician consultant (available in the video review charter). Only the MCL IB1001 study team had access to the randomization sequences. To ensure that central raters are blinded to study subject identifiers, each subject video was assigned a "barcode" and "reading number" that identify the video throughout the central review process. Usage of the "barcode" and "reading number" blinded the rater to any information (i.e., subject ID, DOB, visit identifier) that may introduce bias during central review. |
| Implementation | 10 | Who generated the random allocation sequence, who enrolled participants, and who assigned participants to interventions | The PI was responsible for enrolling all trial participants. No randomization/allocation of participants to different intervention group occurred. |
| Blinding | 11a | If done, who was blinded after assignment to interventions (for example, participants, care providers, those assessing outcomes) and how | 6 |
| | 11b | If relevant, description of the similarity of interventions | All patients received treatment with NALL followed by a post-treatment washout period. |
| Statistical methods | 12a | Statistical methods used to compare groups for primary and secondary outcomes | 7 |
| | 12b | Methods for additional analyses, such as subgroup analyses and adjusted analyses | 7 |
| **Results** | | | |
| Participant flow (a diagram is strongly recommended) | 13a | For each group, the numbers of participants who were randomly assigned, received intended treatment, and were analysed for the primary | 8 |

| | | | |
|---|---|---|---|
| | | outcome | |
| | 13b | For each group, losses and exclusions after randomisation, together with reasons | 8 |
| Recruitment | 14a | Dates defining the periods of recruitment and follow-up | 3 |
| | 14b | Why the trial ended or was stopped | The trial was not ended or stopped prematurely |
| Baseline data | 15 | A table showing baseline demographic and clinical characteristics for each group | 17 |
| Numbers analysed | 16 | For each group, number of participants (denominator) included in each analysis and whether the analysis was by original assigned groups | 20,21 |
| Outcomes and estimation | 17a | For each primary and secondary outcome, results for each group, and the estimated effect size and its precision (such as 95% confidence interval) | 8,9 |
| | 17b | For binary outcomes, presentation of both absolute and relative effect sizes is recommended | N/A |
| Ancillary analyses | 18 | Results of any other analyses performed, including subgroup analyses and adjusted analyses, distinguishing pre-specified from exploratory | 9, 20,21 |
| Harms | 19 | All important harms or unintended effects in each group (for specific guidance see CONSORT for harms) | 9 |
| **Discussion** | | | |
| Limitations | 20 | Trial limitations, addressing sources of potential bias, imprecision, and, if relevant, multiplicity of analyses | 10,11 |
| Generalisability | 21 | Generalisability (external validity, applicability) of the trial findings | 10,11 |
| Interpretation | 22 | Interpretation consistent with results, balancing benefits and harms, and considering other relevant evidence | 10,11 |
| **Other information** | | | |

| | | | |
|---|---|---|---|
| Registration | 23 | Registration number and name of trial registry | 3,5 |
| Protocol | 24 | Where the full trial protocol can be accessed, if available | N/A |
| Funding | 25 | Sources of funding and other support (such as supply of drugs), role of funders | 3,12 |

*We strongly recommend reading this statement in conjunction with the CONSORT 2010 Explanation and Elaboration for important clarifications on all the items. If relevant, we also recommend reading CONSORT extensions for cluster randomised trials, non-inferiority and equivalence trials, non-pharmacological treatments, herbal interventions, and pragmatic trials. Additional extensions are forthcoming: for those and for up to date references relevant to this checklist, see www.consort-statement.org.
